# Optimal upper respiratory tract sampling time for novel coronavirus pneumonia suspects

**DOI:** 10.1101/2020.05.06.20069302

**Authors:** Jing Zhou, Lin Chen, Dehe Zhang, Haijun Chen, Qiyue Sheng, Hongsheng Deng, Yang Zhang, Shunlan Ni, Shengnan Luo, Binbin Ren

**Affiliations:** Department of Infectious Diseases, Jinhua Hospital of Zhejiang University, Jinhua Municipal Central Hospital, Jinhua 32100, Zhejiang Province, China; Department of Intensive Medicine, Jinhua Hospital of Zhejiang University, Jinhua Municipal Central Hospital, Jinhua 32100, Zhejiang Province, China

**Keywords:** COVID-19, nucleic acid detection, exposure, sampling time

## Abstract

**Objective:** Explore best upper respiratory tract sampling time of suspected novel coronavirus pneumonia cases.

**Methods:** We collected dates of patients from Hangzhou, Shenzhen, Jinhua city and so on who had the clear exposure history of a novel coronavirus pneumonia(COVID-19). We retrospected demographic data, exposure time, onset time, visiting time and positive time for novel coronavirus nucleic acid detection in respiratory specimens. There were 256 patients from January 20,2020-February 12,2020 from eight cities included in our study. 106 cases appeared symptoms before January 25^th^ and 150 after.

**Results:** There were 136(53.1%)male infected cases. The mean age of all patients was 43.80±14.85. The median time from exposure to onset was 5(3,8) days. The median time of the first time of positive nucleic acid detection was 11(9,14)days and mode number was 13. The median time from onset to the first time of positive nucleic acid detection was 6(4,8)days and mode number was 5. The time from onset to definite diagnosis was 5(3,7) days before January 25^th^ while it was 7.5(5,10)days after which was significantly shorter before January 25^th^(U=3885.5,P<0.001). The time from exposure to definite diagnosis was 11(9,14)days and 11(9,14)days before January 25^th^ and after and without significant difference. The time from exposure to definite diagnosis was 11(9,13)days in first-tier cities and 13(11,15)days in second and third-tier cities. The difference was significantly shorter of first-tier cities(U=1355.5, P=0.039). And also the time was short from visiting to definite diagnosis which was 2(2,3)days in first-tier cities and 3(2,4)days in second and third-tier cities but without significant difference(U=842.5, P=0.054).

**Conclusions:** From our study we found that the best upper respiratory tract sampling time for novel coronavirus pneumonia suspects was 13days after exposure. The time from onset to definite diagnosis was shorter after January 25^th^. The patients were diagnosed faster in the first-tier cities after exposure.

## Introductions

Novel coronavirus infected patients with pneumonia have been discovered in Wuhan, Hubei since at the end of the December 2019. The epidemic has spread all of the the country and the wold make a great threat to the public health. There were 114 countries had infected patients and 118 000 patients in total until March 11^th^ with a mortality of 3.6%. The first sequencing of the viral RNA gene was completed in January 3rd by Chinese microbiologists and medical experts. The CDC shared the new gene sequence of the coronavirus to the world in January 10^th^. They developed and tested PCR diagnostic reagent which was wildly used in clinic since January 11^th^ [1]. The novel coronavirus pneumonia was caused by novel coronavirus (2019-nCOV) which was renamed as SARS-CoV-2 in February 11^th^ by the the International Committee on Taxonomy of Viruses. SARS-CoV-2 belongs to the novel coronavirus of beta genus, with capsule, round or oval, 60-140nm in diameter and positive RNA virus. In vitro, sars-cov-2 can be found in human respiratory epithelial cells in about 96 hours [2]. Early novel coronavirus pneumonia patients had a history of Wuhan residence or contacted with the people from Wuhan, suggesting that there was interpersonal transmission [3,4]. Covid-19 is a highly infectious disease. The R0 of covid-19 in early literature is 2.2 ~ 2.9 [1,5] and 3.77 in another study including 8 866 patients [6]. Therefore, early diagnosis, isolation and treatment of suspected cases are essential to control the epidemic. The detection of novel coronavirus nucleic acid is an effective method for rapid diagnosis [7,8]. However when is the optimal upper respiratory tract sampling time is not sure. The optimal sampling time can help us to diagnosis in time at the same time to avoid the waste of the medical resources.

In this study, we analyzed the distribution of the first positive time after exposure from different cities in order to obtain the optimal optimal upper respiratory tract sampling time.

## 1. Object and method

### 1.1 research object

There were 256 patients with clear exposure history of a novel coronavirus pneumonia(COVID-19) included. We collected exposure time, onset time, visiting time of some patients and the first time of respiratory tract specimen to be confirmed positive of novel coronavirus nucleic acid detection.

### 1.2 Research method

### 1.2.1 Diagnostic criteria

COVID-19 was diagnosed by novel coronavirus pneumonia diagnosis and treatment plan issued by the People’s Republic of China health and Health Committee (trial version sixth). All the suspected patients received the test of the samples of sputum, throat swab or secretion of lower respiratory tract by real-time fluorescence quantitative PCR to show that sars-cov-2 nucleic acid was positive.

### 1.2.2 Data collection

The data was collected and obtained from the electronic case system which was came from official website of the Municipal Health Committee and the mainstream media of the city. All the data collected were reviewed by senior doctors in order to insure the accuracy.

### 1.2.3 Inclusion criteria

The patients were consistent with the diagnostic criteria and the following conditions: a: a clear history of Epidemiology (defined contact initiation and end time), b: single exposure, c: exposure time not exceeding 1 day. Exclusion criteria: a: history of multiple exposures. B: only one exposure but the time (from contact initiation to end time) was more than one day. c: without an unclear exposure time.

### 1.2.4 Grouping method

The patients were divided into the group before January 25^th^ including 106 patients and 150 patients in group after January 25^th^. The patients with the confirmed visiting time in fist tier city group was 72 cases and 89 cases in second tier and third tier city group.

Statistical analysis: Statistical analyses were performed using SPSS 23.0 (International Business Machines Corporation, IBM, USA). Normally distributed. Normally distributed continuous variables are summarized as the mean and standard deviation and t-test was applied to test differences between two groups; Non-normally distributed data are recorded as median and interquartile range (Q25, Q75) as appropriate., and U-test was applied to test differences between two groups; P < 0.05 was statistically significant.

## 2. Result

### 2.1 General information

There were 136 (53.1%) cases were male and 120(46.9%) female, the average age was 43.80 ± 14.85 years old, most of them were between 31 and 70 years old (80.9%). (Figure 1.)

**Figure 1.**
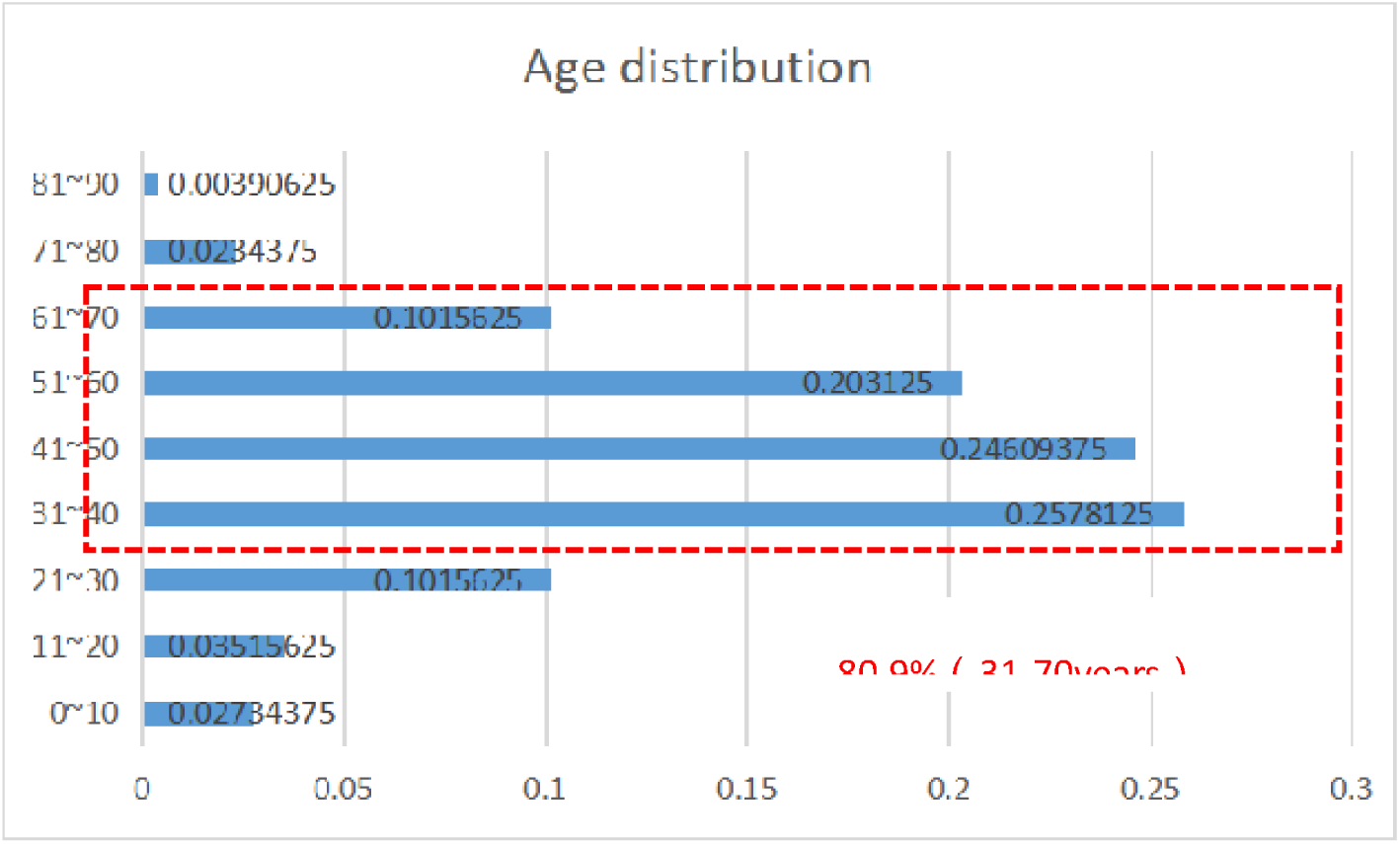
Age distribution of COVID-19 patients

### 2.2 Time information

The median incubation period was 5(3,8)days and the time range was 0-18days. The median time from onset to the first time of confirmed nucleic acid positive was 5(4,8)days. The median time from exposure to the first time confirmed nucleic acid positive was 11(9,14)days with mode number 13day (Figure 2, table 1)

**Figure 2.**
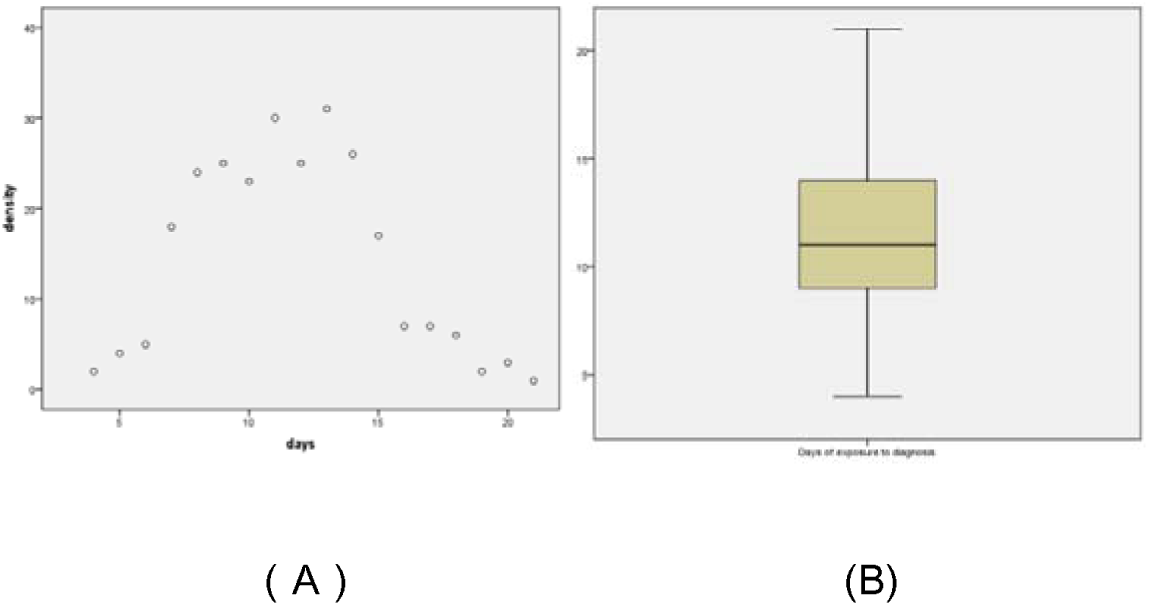
Time distribution of nucleic acid positive from exposure to first swab of pharynx(A is a scatterplot, B is a boxplot)

**Table 1.**
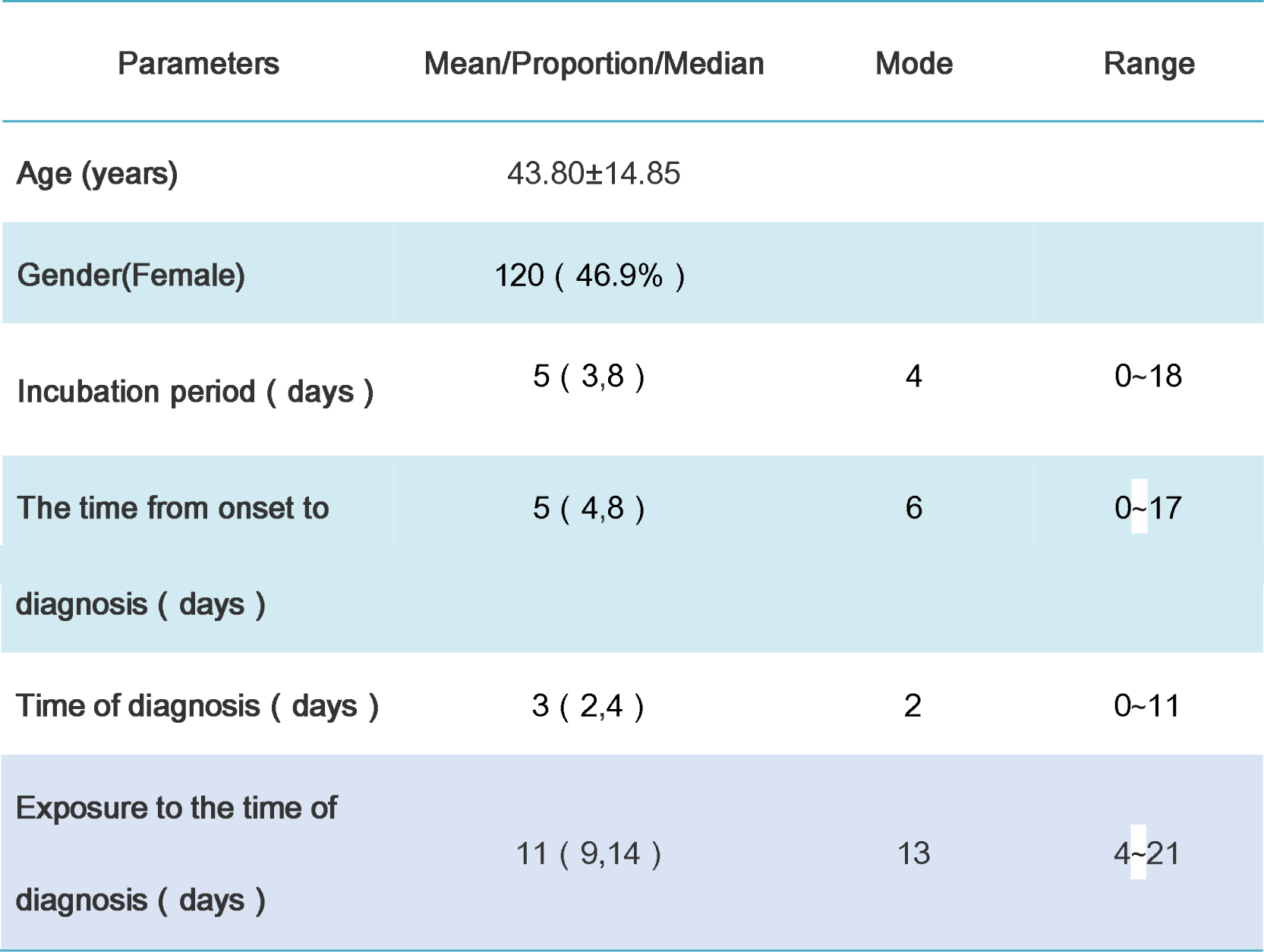
Demographic characteristics and characteristics of each time period after exposure

### 2.3 Comparison onset time

There was no significant difference between the two groups in age, gender and the time from exposure to diagnosis between patients before January 25^th^ and after. But the median time from onset to diagnosis was 5(3,7) days before January 25^th^ which was significantly shorter than 7.5(5,10) days(U=3885.5, P<0.001) (Table 2).

**Table 2.**
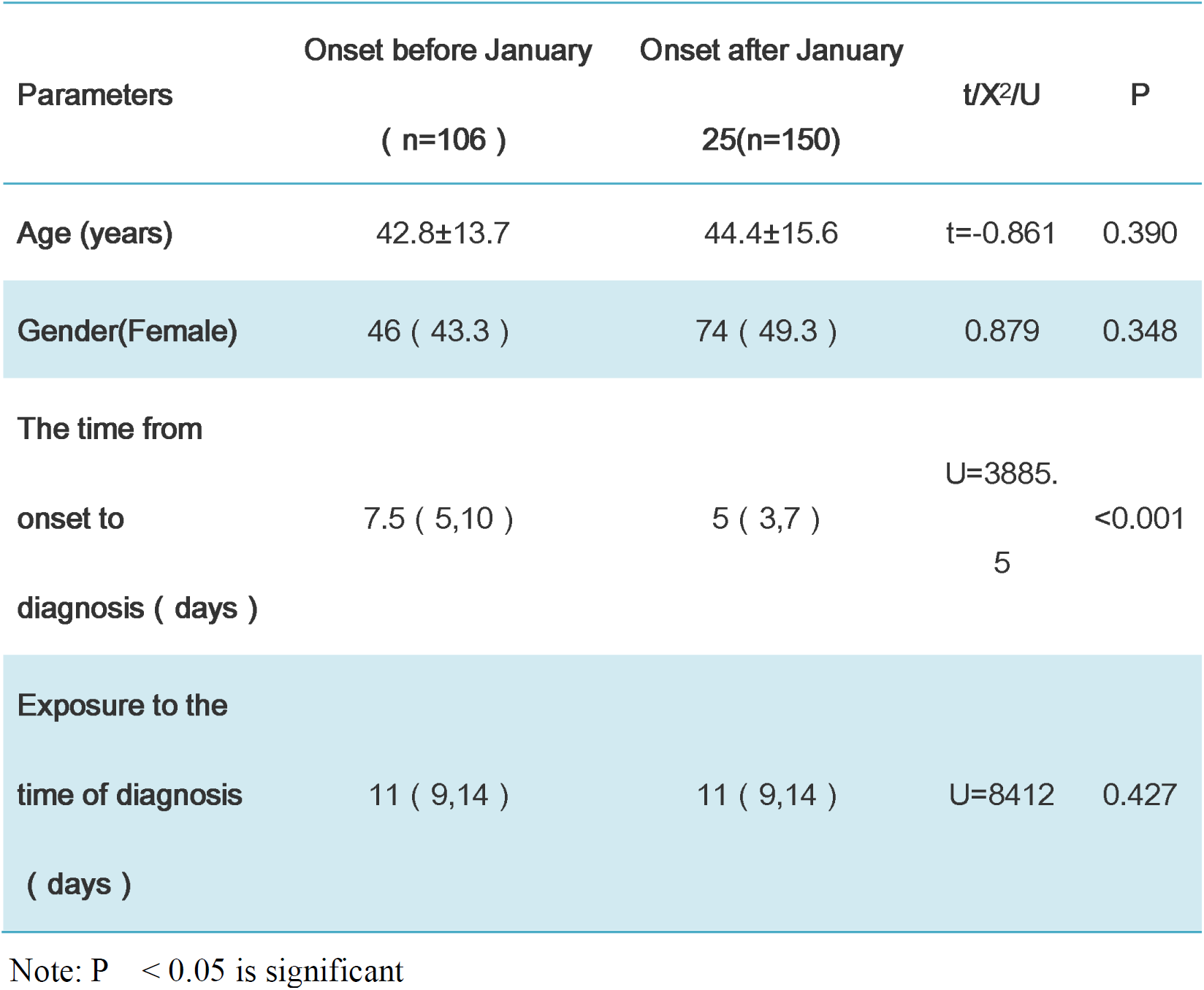
Comparison between the group with pre-january 25 onset and the group with post-january 25 onset

| Parameters | Onset before January<br>( n=106 ) | Onset after January<br>25(n=150) | t/X<sup>2</sup>/U | P |
| --- | --- | --- | --- | --- |
| Age (years) | 42.8±13.7 | 44.4±15.6 | t=-0.861 | 0.390 |
| Gender(Female) | 46 ( 43.3 ) | 74 ( 49.3 ) | 0.879 | 0.348 |
| The time from<br>onset to<br>diagnosis ( days ) | 7.5 ( 5,10 ) | 5 ( 3,7 ) | U=3885.5 | <0.001 |
| Exposure to the<br>time of diagnosis<br>( days ) | 11 ( 9,14 ) | 11 ( 9,14 ) | U=8412 | 0.427 |

Note: P < 0.05 is significant

### 2.4 Comparison different regions

The age of onset was younger and the proportion of females was lager in the first tier cities. The median time from exposure to diagnosis was 11(9,13)days which was significantly shorter than 13(11,15)days in the second and third tier cities group(U = 1355.5, P = 0.039). The median time from visiting to diagnosis was 2(2,3) days which was fast than 3(2,4)days in the second and third tier cities but without significant difference(U=842.5, P=0.054) (Table 3).

**Table 3.** Comparison between the first-tier city group and the second - and third-tier city group

| Parameters | First-tier cities<br>( n=72 ) | Second - and<br>third-tier cities<br>(n=89) | t/X<sup>2</sup>/U | P |
| --- | --- | --- | --- | --- |
| Age (years) | 41.39±16.04 | 46.75±16.07 | t=2.108 | 0.037 |
| Gender(Female) | 39 ( 54.1 ) | 36 ( 40.4 ) | 3.010 | 0.083 |
| Time of diagnosis<br>( days ) | 2 ( 2,3 ) | 3 ( 2,4 ) | U=842.5 | 0.054 |
| Exposure to the<br>time of diagnosis<br>( days ) | 11 ( 9,13 ) | 13 ( 13,15 ) | U=1355.5 | 0.039 |

Note: P < 0.05 is significant

## Discussion

Novel coronavirus pneumonia is infectious at the early stage of onset. SARS-CoV-2 can be found in human respiratory epithelial cells after infected about 96 hours when isolated and cultured in vitro [2]. The load of viral nucleic acid reached the peak within 2-5 days after the onset of the disease while the load of previous SARS CoV viral reached the peak about 10 days after the symptoms appeared [9]. Real time fluorescent RT-PCR is the most commonly used diagnostic method at present which amplify the specific nucleic acid sequence in the specimen. After more than 40 times of amplification, the quantity of nucleic acid can reach enough to be detected by conventional methods such as fluorescence.

In this study, 136(52.9%) male patients were more than female which might be the reason that expression of ACE2 was higher in Asian men [10,11]. There were 80.9% patients distributed from 31 to 70 years old which was similar to the study of the center of Disease Control and Prevention [12].

The incubation period of infectious diseases refers to the time from the entry of pathogenic microorganisms into the human body to the first symptoms of infection, which is an important basis for the isolation time limit of close contacts. The incubation period of new coronavirus is usually 1-14 days longer than that of SARS CoV for 2-10 days and mostly 3-7 days. In our study the incubation period was 0~18d, median time was 5(3,8)days, mean time was 5.5days and mode number was 4days which was similar as Li reported the retrospective research of the early stage of the epidemic including 425 patients [1]. However the cases involved in our study were all single exposure with a time span no more than 1day. There were 161 cases with confirmed visiting time, though the analysis of these patients the median time from visiting to diagnosis was 3(2,4)days and mode number was 2days. Although the time from visiting to diagnosis was in lager span time from 0 to 11days, only 7 cases were more than 6days, mainly concentrated in 0-6days. The reason for delay diagnosis may be due to the shortage of medical resources in some hospitals, on the other hand the early diagnosis of the disease should be confirmed by the expert of Health Commission. And finally the false negative of nucleic acid test of respiratory could also cause the delay diagnosis.

The time from exposure to diagnosis of 256 patients was from 4 to 21 days, the median time was 11(9,14)days and mode number was 13 days. There were also several mainly reasons. First of all, the patients would not actively to see the doctor because of no evident symptoms during incubation period. And then due to the sensitivity of nucleic acid detection and sample quality the positive rate of pharyngeal sample nucleic acid detection might be less than 50% [13]. The positive rate may be higher of the sample form lower respiratory tract(sputum and bronchoalveolar lavage). However at the early stage of the disease the lesion is usually located in the lateral zone of the lung with the presentation of dry cough which cause the difficulty to obtain the sample form lower respiratory tract. At the early stage of infection the lower load of viral from upper respiratory tract was another reason leads to false negative test.

We divided groups into before January 25^th^ and after because the first level response of major public health event was launched successively in most regions of the country from January 23^th^ to 25^th^. We found that the median time from exposure to diagnosis was no significance between two groups but the median time from onset to diagnosis was significantly longer before January 25^th^ group. It was mainly because of the rapid response of the Chinese government to take many strategies including publish epidemic news, providing enough medical resources and so on. In such case, the patient would go to hospital as soon as they had the symptoms.

## Conclusion

The median time from onset to diagnosis was significantly shorter after January 25th group. The optimal upper respiratory tract sampling time from our study was 13 days after exposure. However the sample of our study was small, further larger sample is needed.

## Data Availability

The data used to support the findings of this study are available from the corresponding author upon request

## Acknowledgments

This work was supported by the Jinhua Science and Technology Bureau (2020XG-06).

## Ethics approval and consent to participate

The ethics committee of the hospital approved the study

## References

[1] Li Q, Guan X, Wu P, et al. Early Transmission Dynamics in Wuhan, China, of Novel Coronavirus-Infected Pneumonia[J]. The New England journal of medicine, 2020:1–9.DOI:10.1056/NEJMoa2001316.

[2] Zhu N, Zhang D, Wang W, et al. A Novel Coronavirus from Patients with Pneumonia in China, 2019[J]. N Engl J Med, 2020,382(8):727–733. DOI:10.1056/NEJMoa2001017.

[3] Chan J F, Yuan S, Kok K H, et al. A familial cluster of pneumonia associated with the 2019 novel coronavirus indicating person-to-person transmission: a study of a family cluster[J]. Lancet, 2020,395(10223):514–523. DOI:10.1016/S0140-6736(20)30154-9.

[4] Yu P, Zhu J, Zhang Z, et al. A familial cluster of infection associated with the 2019 novel coronavirus indicating potential person-to-person transmission during the incubation period[J]. J Infect Dis, 2020.DOI:10.1093/infdis/jiaa077[publishedOnlineFirst:2020/02/18]

[5] Tao Liu J H J X, Guanhao H M K Z, Lifeng, et al. Time-varying transmission dynamics of Novel Coronavirus Pneumonia in China[J]. bioRxiv, 2020. DOI: https://doi.org/10.1101/2020.01.25.919787.

[6] Yang Yang Q L, Ming-Jin L, Yi-Xing W, et al. Epidemiological and clinical features of the 2019 novel coronavirus outbreak in China[J]. medRxiv, 2020.DOI: https://doi.org/10.1101/2020.02.10.20021675.

[7] Chu D, Pan Y, Cheng S, et al. Molecular Diagnosis of a Novel Coronavirus (2019-nCoV) Causing an Outbreak of Pneumonia[J]. Clin Chem, 2020:1–15. DOI:10.1093/clinchem/hvaa029[publishedOnlineFirst:2020/01/31]

[8] Corman V M, Landt O, Kaiser M, et al. Detection of 2019 novel coronavirus (2019-nCoV) by real-time RT-PCR[J]. Euro Surveill, 2020,25(3):1–8. DOI:10.2807/1560-7917.ES.2020.25.3.2000045.

[9] Zou L, Ruan F, Huang M, et al. SARS-CoV-2 Viral Load in Upper Respiratory Specimens of Infected Patients[J]. N Engl J Med, 2020. DOI:10.1056/NEJMc2001737[publishedOnlineFirst:2020/02/19]

[10] Cao Y, Li L, Feng Z, et al. Comparative genetic analysis of the novel coronavirus (2019-nCoV/SARS-CoV-2) receptor ACE2 in different populations[J]. Cell Discov, 2020, 6:11. DOI:10.1038/s41421-020-0147-1[publishedOnlineFirst:2020/02/24]

[11] Zhao Y, Zixian Z, Yujia W, et al. Single-cell RNA expression profiling of ACE2, the putative receptor of Wuhan 2019-nCov[J]. bioRxiv, 2020. DOI:10.1101/2020.01.26.919985.

[12] Response E W G F. [The epidemiological characteristics of an outbreak of 2019 novel coronavirus diseases (COVID-19) in China][J]. Zhonghua Liu Xing Bing Xue Za Zhi, 2020, 41(2):145–151. DOI:10.3760/cma.j.issn.0254-6450.2020.02.003.

[13] Gao Z C. [Efficient management of novel coronavirus pneumonia by efficient prevention and control in scientific manner][J]. Zhonghua Jie He He Hu Xi Za Zhi, 2020, 43(3):163–167. DOI:10.3760/cma.j.issn.1001-0939.2020.0001.

